# Thyroid Function Analysis in COVID-19: A Retrospective Study from a Single Center

**DOI:** 10.1101/2021.02.09.21251435

**Authors:** Jahanzeb Malik, Asmara Malik, Muhammad Javaid, Tayyaba Zahid, Uzma Ishaq, Muhammad Shoaib

**Affiliations:** Department of Cardiology, Rawalpindi Institute of Cardiology, Rawal Road, Rawalpindi, 46000, Pakistan; Department of Public Health, National University of Medical Sciences, Mall road, Saddar, Rawalpindi, 46000, Pakistan; Department of Pathology, Foundation University Medical College, Islamabad, 44000, Pakistan; Department of Cardiology, Pakistan Institute of Medical Sciences, Islamabad, 44000, Pakistan

**Keywords:** COVID-19, Thyroid diseases, Thyroid-Stimulating Hormone, Triiodothyronine

## Abstract

**Background and Objective:** Coronavirus disease 2019 (COVID-19) is an on-going epidemic with a multitude of long-ranging effects on the physiological balance of the human body. It can cause several effects on thyroid functions as well. We aimed to assess the lasting sequelae of COVID-19 on thyroid hormone and the clinical course of the disease as a result.

**Methods:** Out of 76 patients, 48 patients of COVID-19 positive and 28 patients of COVID-19 negative polymerase chain reaction (PCR) were assessed for thyroid functions, IL-6, and Procalcitonin between moderate, severe, and critical pneumonia on HRCT.

**Results:** Seventy-five percent of patients with COVID-19 had thyroid abnormalities and higher IL-6 levels (76.10 ± 82.35 vs. 6.99 ± 3.99, 95% CI 52.18-100.01, P-value <0.01). Logistic regression analysis suggested TT3 (P-value 0.01), IL-6 (P-value <0.01), and Procalcitonin (P-value 0.03) as independent risk factors for COVID-19. ROC curve demonstrated IL-6 as the most sensitive marker (P-value <0.01), and TT3, and Procalcitonin as the predictor for COVID-19 disease.

**Conclusion:** This pilot study from Pakistan demonstrates that changes in serum TSH and TT3 levels may be important manifestations of the courses of COVID-19 pneumonia.

## Introduction

The on-going pandemic of coronavirus disease 2019 (COVID-19) has been found to have multiple long-ranging effects on the normal physiological balance of the human body. Complex and severe effects on human organ systems are being identified rapidly, including the respiratory, gastric, circulatory, immune, renal, hepatic, cardiac, and hematological systems. Literature on COVID-19 affecting human thyroid function is increasing gradually and the understanding of thyroid dysfunction and its mechanism is growing. The expression of ACE2 combined with transmembrane protease serine 2 (TMPRESS2) is the key cellular complex for the virus to infect the human cells and interestingly both expression levels are present in the thyroid gland, even more than the lungs.^1^

COVID-19 initiates an immune response over-activity leading to release of pro-inflammatory cytokines, particularly interleukin-6 (IL-6), which leads to overt thyroid dysfunction by disruption of desiodases and thyroid transport proteins. Levels of T3 is inversely proportional to IL-6 with a modest decrease of TSH and T4. This abnormality is described as sick euthyroid syndrome.^2,3^ During the course of COVID-19 pneumonia, thyrotoxicosis may be caused secondary to graves thyroiditis or subacute inflammatory thyroiditis. It can be complicated by arrhythmias or thromboembolic episodes. Hence, abnormalities of thyroid dysfunction are important to evaluate in COVID-19.^4^

We conducted a retrospective, cross-sectional study of COVID-19 patients in our hospital in Rawalpindi, Pakistan, compared with a matched cohort of non-COVID-19 patients. Our objectives were to determine if COVID-19 caused acute and subacute thyroid abnormalities, as determined by a diagnostic assessment of thyroid function, and whether thyroid dysfunction leads to a more severe clinical course of COVID-19 in a South Asian population.

## Materials and Methods

This retrospective study was approved by the ethical review board of Foundation University (FFH/37/DCA/2020) following the World Medical Association Declaration of Helsinki. Informed written consent was taken from all the participants. Patients were considered for inclusion with positive high resolution computed tomography (HRCT) for SARS-CoV-2. HRCT was considered positive if it had consolidation, linear opacity, septal thickening, crazy-paving pattern, or halo sign. Positive COVID-19 polymerase chain reaction (PCR) was labelled group1 while negative PCR were labelled group 2.

The medical records of all the patients admitted at our institute from April 2020 to July 2020 with pneumonia were retrospectively examined. All records were accessed from October 2020. Patients with a history of thyroid disease, without assessment and follow-up of thyroid functions, IL-6 and procalcitonin, pregnancy, and mild COVID-19 infections were excluded from the study. A total of 76 patients were enrolled in this study. Forty-eight patients were enrolled in group 1 and 28 in group 2. All patients had their tests on day one before initiation of treatment and after three months’ follow-up. This group was screened for underlying thyroid disease or any other medical history that can affect thyroid function.

All the cases were labeled as moderate, severe, or critical based on the clinical symptoms, laboratory tests, and chest computed tomography (CT) scans. Moderate and severe cases were classified as having pneumonia manifestations seen on imaging, respiratory rate of ≥ 30 breaths/min, and oxygen saturation ≤ 90% at rest. Critical cases were labeled as those requiring mechanical ventilation due to respiratory failure, other organ failures that require intensive care unit, and the presence of shock.

All patients included in the study had their thyroid functions, including thyroid-stimulating hormone (TSH, reference range: 0.45 – 4.5 mIU/L), total triiodothyronine (TT3, reference range: 70 to 200 ng/dL), serum total thyroxine (TT4, reference range: 4.6 – 11.2 mcg/dL), procalcitonin (reference range: < 0.05 mcg/L), and IL-6 (reference range: 0 −16.4 pg/mL) levels done via electrochemiluminescent immunoassay (ECLIA) in the Elecsys® 2010 immunoassay system. All the parameters were analyzed and compared between the COVID-19 and the control group. Any deviation from normal reference range was considered as thyroid dysfunction.

Data were analyzed with the Statistical Package for the Social Sciences (SPSS) version 26 (IBM Corp., Armonk, NY, USA.). Quantitative data were presented as mean ± standard deviation (SD) and qualitative data in frequency and percentages. Both groups were compared using the students’ t-test and Chi-square when feasible. A receiver operating characteristic (ROC) curve was calculated for thyroid function tests and the acute phase reactants to find out sensitivity and specificity of the lab parameters in COVID-19 pneumonia. Confidence interval (CI) were presented for the lab parameters in all groups. A P-value of less than 0.05 was considered significant.

## Results

During the course of our study, a total of 76 patients were enrolled. Forty-eight with confirmed COVID-19 and 28 patients with non-COVID-19 pneumonia with their thyroid profiles done were also included as controls. Patient demographics and characteristics of pneumonia in both the groups were evenly matched. They are differentiated in **Table 1**.

**Table 1.** Demographics and severity of pneumonia

| Variable | COVID-19(n=48) | Non-COVID-19<br>(n=28) | P-value |
| --- | --- | --- | --- |
| Age (Mean ± SD) | 51 ± 19.30 | 64.79 ± 11.44 | 0.68 |
| Male (%) | 31(64.6%) | 15(53.6%) | 0.34 |
| Severity of pneumonia |  |  |  |
| Moderate | 22(45.8%) | 8(28.6%) | 0.13 |
| Severe | 9(18.8%) | 10(35.7%) | 0.09 |
| Critical | 17(35.4%) | 10(35.7%) | 0.97 |

Our examination of the thyroid function analysis suggested that 36 (75%) patients with COVID-19 pneumonia had abnormal thyroid functions while 24 (85.7%) patients were detected with non-COVID-19 pneumonia. IL-6 was significantly higher in COVID-19 group as compared with non-COVID-19 group (76.10 ± 82.35 vs. 6.99 ± 3.99, 95% CI 52.18-100.01, P-value <0.01).

At follow-up, there was a downtrend in IL-6 levels in both groups (14.16 ± 16.58 vs. 4.88 ± 2.66, P-value <0.01). Procalcitonin levels were statistically significant between both the groups (P-value <0.01). IL-6 was associated with abnormal TSH, TT3, and TT4 levels (P-value <0.01). **Table 2** differentiates thyroid profile and acute phase reactants in both COVID-19 and non-COVID-19 group.

**Table 2.** Comparison of thyroid functions and acute phase reactants in both groups

| Tests | COVID<br>(n=48) |  | 95% CI | Non-COVID<br>(n=28) |  | 95% CI | P-value |
| --- | --- | --- | --- | --- | --- | --- | --- |
|  | At<br>admission | At<br>Follow-up |  | At<br>admission | At<br>Follow-up |  |  |
| **IL6**<br>**(pg/mL)** | $76.10 \pm 82.35$ | $14.16 \pm 16.58$ | 52.18-100.01 | $6.99 \pm 3.99$ | $4.88 \pm 2.66$ | 5.44-8.54 | $<0.01$ |
| **Procalcitonin (mcg/L)** | $0.36 \pm 0.52$ | $0.14 \pm 0.32$ | 0.21-0.51 | $0.06 \pm 0.12$ | $0.05 \pm 0.08$ | 0.01-0.11 | $<0.01$ |
| **TSH**<br>**(mIU/L)** | $1.48 \pm 2.47$ | $1.11 \pm 1.15$ | 0.76-2.20 | $1.73 \pm 2.06$ | $1.65 \pm 1.73$ | 0.93-2.53 | $<0.01$ |
| **TT3**<br>**(ng/dL)** | 74.62 ±<br>33.71 | 89.98 ±<br>35.15 | 64.83-84.42 | 100.21 ±<br>45.46 | 101.86<br>±<br>40.90 | 82.59-117.84 | <0.01 |
| **TT4**<br>**(mcg/dL)** | 6.67 ±<br>2.74 | 6.79 ±<br>2.37 | 5.87-7.46 | 6.43 ±<br>2.44 | 9 ±<br>2.77 | 5.48-7.38 | 0.11 |

Comparison of TSH and TT3 shows significantly lower mean values in severe COVID-19 infection as compared with critical patients (0.32 ± 0.22 vs. 2.32 ± 3.44, P-value <0.01) (67.44 ± 34.51 vs. 77.12 ± 48.22, P-value <0.01). A comparison of lab parameters among different classes of pneumonia is shown in **Table 3**.

**Table 3.**
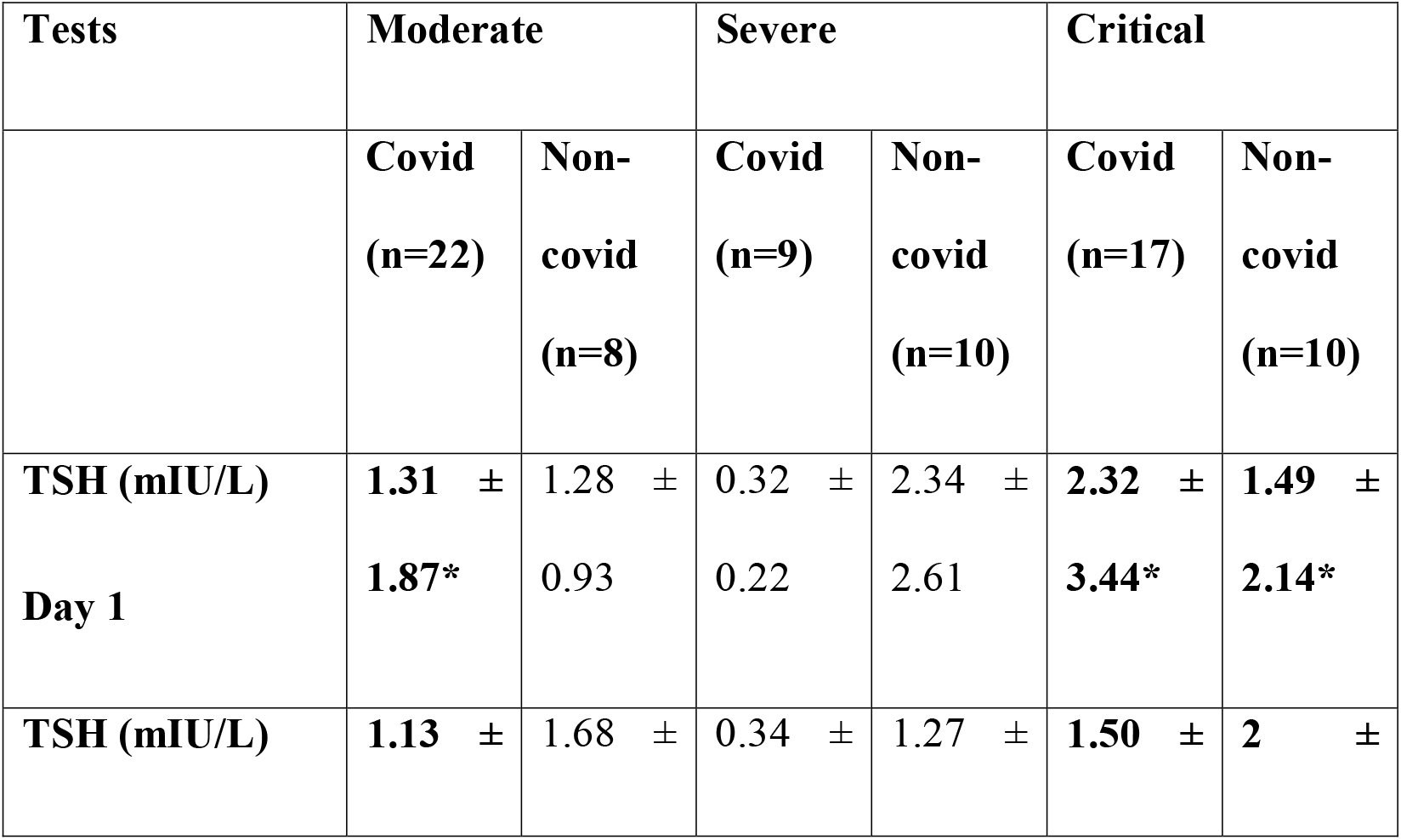
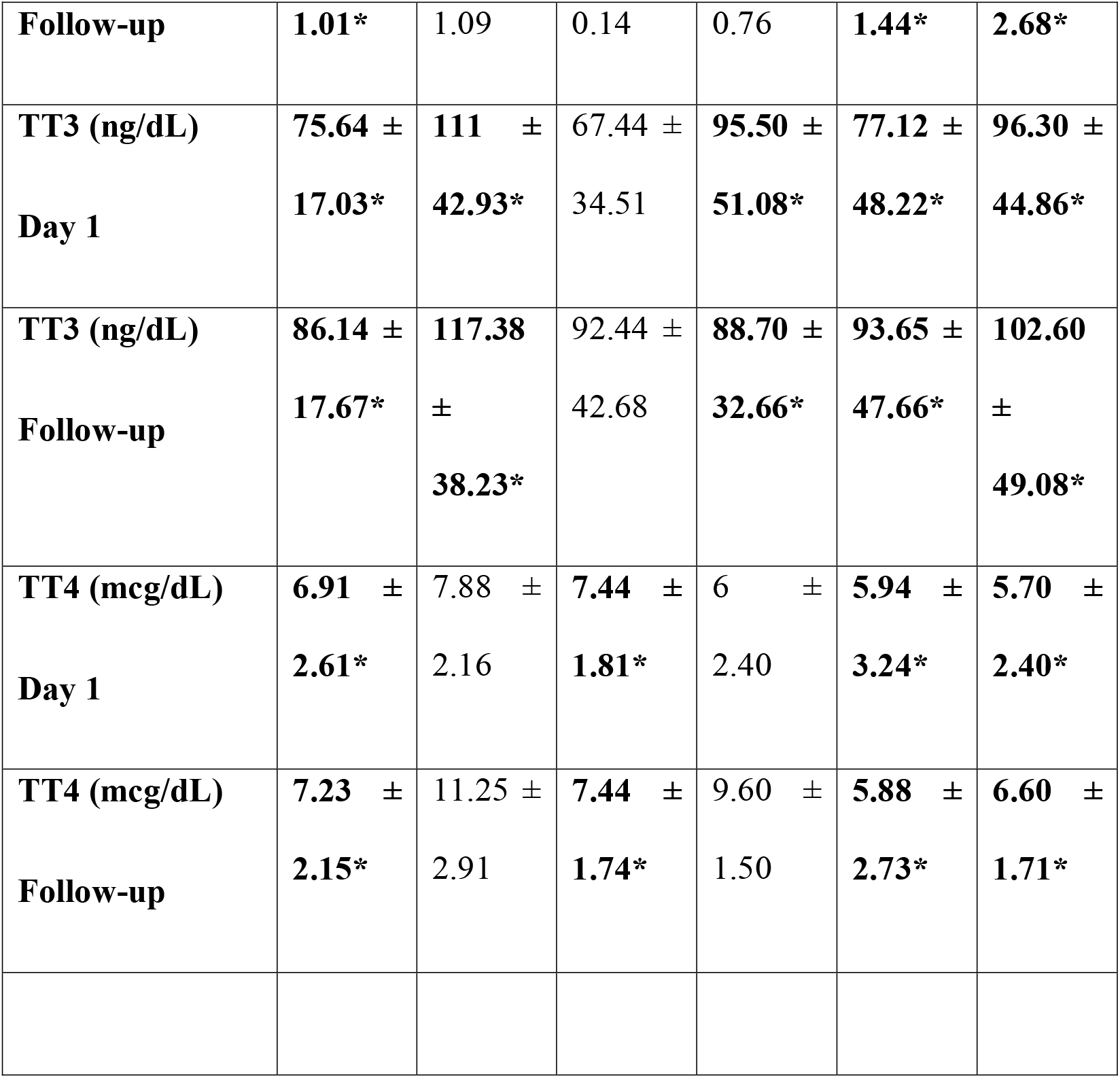
Comparison of thyroid funcitons between severity of pneumonia at day 1 and at follow-up. **\*P-value <0.05**

**Table 4.** Sensitivity and specificity in predicting COVID-19

| Test | Area under the curve | Sensitivity | Specificity | P-value |
| --- | --- | --- | --- | --- |
| IL6 | 0.941 | 0.979 | 0.357 | <0.01 |
| Procalcitonin | 0.745 | 0.667 | 0.357 | <0.01 |
| TSH | 0.378 | 0.438 | 0.679 | 0.07 |
| TT3 | 0.283 | 0.375 | 0.786 | <0.01 |
| TT4 | 0.542 | 0.479 | 0.393 | 0.54 |

**Table 5.** Multivariate analysis of lab parameters in moderate, severe, and critical pneumonia

| Moderate Pneumonia |  |  |  |  |  |
| --- | --- | --- | --- | --- | --- |
| Lab Parameters | B | SE | Wald | OR, 95% CI | P-value |
| IL-6 | 0.02 | .01 | 3.49 | 1.67, 10.46-25.00 | 0.06 |
| TT3 | 0.00 | .00 | 0.64 | 0.12, 73.81-96.32 | 0.42 |
| **TT4** | -0.24 | .12 | 4.20 | 23.81, 6.23-8.10 | 0.04 |
| **TSH** | 0.14 | .14 | 0.91 | 0.67, 0.69-1.92 | 0.33 |
| **Procalcitonin** | 2.73 | 1.73 | 2.49 | 0.15, 0.28-0.12 | 0.11 |
| **Severe Pneumonia** |  |  |  |  |  |
| **IL-6** | 0.00 | 0.00 | 1.53 | 0.32, 16.32-48.60 | 0.21 |
| **TT3** | 0.00 | 0.00 | 0.18 | 0.76, 60.43-103.99 | 0.66 |
| **TT4** | -0.01 | 0.10 | 0.01 | 0.35, 5.62-7.75 | 0.90 |
| **TSH** | 0.06 | 0.13 | 0.21 | 0.41, 0.35-2.40 | 0.64 |
| **Procalcitonin** | -0.27 | 0.68 | 0.15 | 0.39, 0.01-0.47 | 0.69 |
| **Critical Pneumonia** |  |  |  |  |  |
| **IL-6** | -0.02 | 0.00 | 7.80 | 1.83, 59.83-140.15 | 0.00 |
| **TT3** | -0.00 | 0.00 | 1.01 | 0.76, 65.60-102.85 | 0.01 |
| **TT4** | 0.29 | 0.13 | 5.29 | 0.91, 4.70-7.01 | 0.02 |
| **TSH** | -0.17 | 0.12 | 1.78 | 0.16, 0.82-3.20 | 0.18 |
| **Procalcitonin** | -1.17 | 0.77 | 2.26 | 2.53, 0.23-0.67 | 0.13 |

Logistic regression was performed to determine how levels of thyroid functions and phase reactants affect the risk of COVID-19 pneumonia. The model suggested TT3 (P-value 0.01), IL-6 (P-value <0.01), and Procalcitonin (P-value 0.03) as independent risk factors for COVID-19. None of the patients received thyroid hormone replacement therapy, and the levels of all the thyroid hormones returned to normal after recovery.

The stem and leaf plot was generated to demonstrate the distribution of the lab parameters. The area under the ROC (AUC) for IL-6 and Procalcitonin was 0.941 (95% CI 0.89-0.98, P-value <0.01) and 0.745 (95% CI 0.00-0.63, P-value <0.01), respectively. TT3 was a predictor of COVID-19 pneumonia with AUC 0.283 (95% CI 0.15-0.40, P-value <0.01**). Figure 1** suggests that thyroid function tests are not good predictors for progression to severe COVID-19 infection. The ROC curve indicated IL-6 to be inversely proportional to TT3.

**Figure 1.**
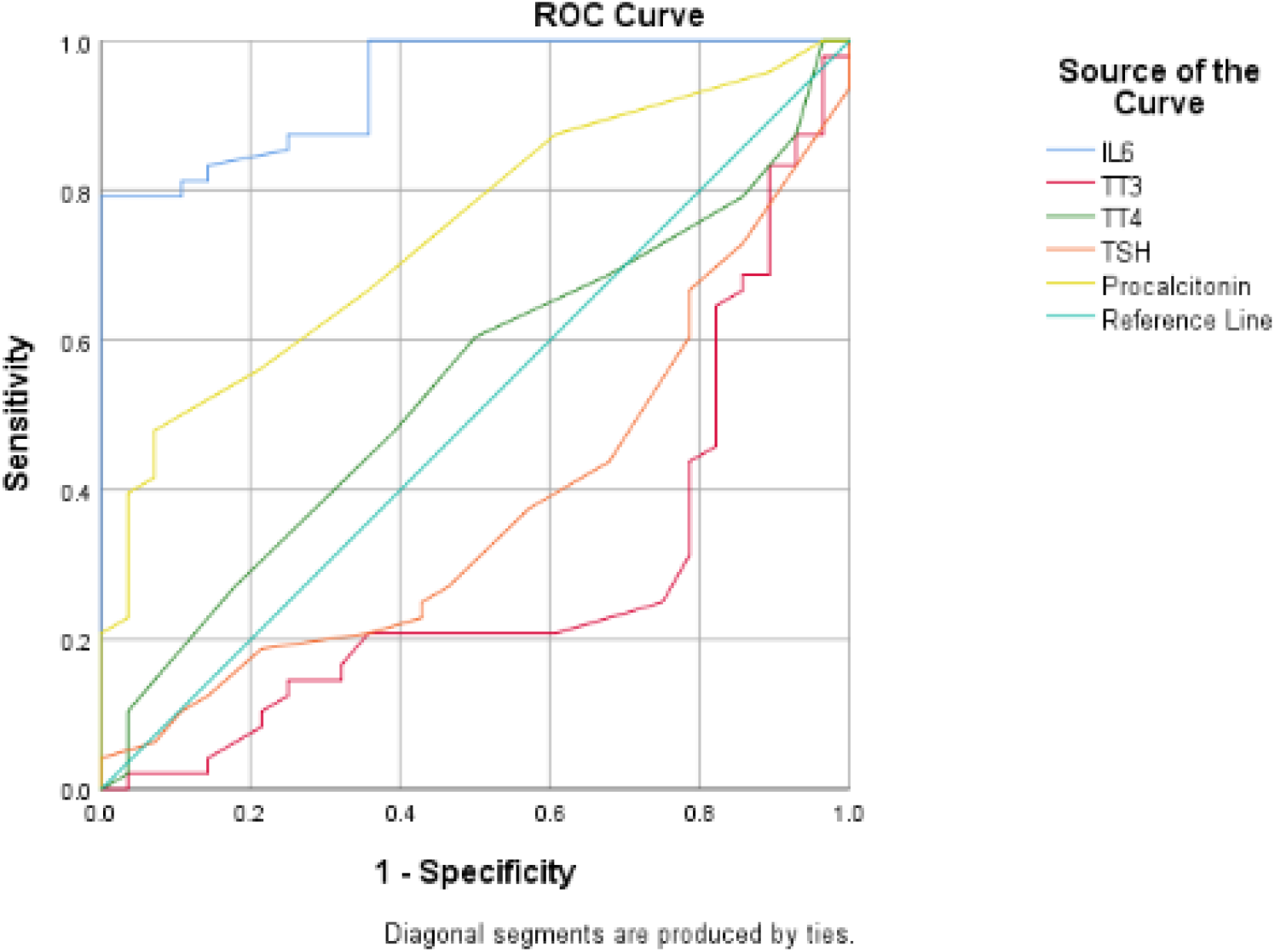
ROC curve for lab parameters. Differentiation of sensitivity and specificity is shown in **Table 4** and Multivariate analysis in **Table 5.**

## Discussion

The results of our study showed that TSH was raised in moderately and critically affected patients of COVID-19 as compared to non-COVID-19 patients. Additionally, TT3 levels were raised significantly more at follow-up in COVID-19 patients than non-COVID-19 patients with little to no discernable effect on TT4 levels. Our findings are in contrast to a similar retrospective study from China, which found lower TSH levels in more severely affected patients of COVID-19 and a study from Italy in which COVID-19 patients were found to have thyrotoxicosis after a confirmatory diagnosis of COVID-19.^5,6^ Although all thyroid function tests returned close to baseline at follow-up in both the Chinese and Italian studies, our study sample showed an outlier in having raised TT3 levels across all three levels of severity of COVID-19. This is of great concern, particularly when paired with the lasting damage to cardiac and respiratory systems which has been documented with regards to COVID-19.^7^ Cardiac abnormalities secondary to thyroid abnormalities may worsen when regarded in the context of post-infectious sequelae.

The thyroid gland is a major organ in maintaining many long term functions of the body, in particular, cardiovascular, respiratory, and catabolic tasks, which is why many lower-income countries have implemented salt iodination as a way to counter the consequences of iodine-deficient thyroid disorders.^8,9^ As an emerging disease, COVID-19 has taken a devastating toll in lower-middle-income countries, with seemingly no end in sight to the rising death toll which, at the time of this writing, stands to cross more than 1, 000, 000.^10^ As the pandemic rages on, more and more insights are emerging into not just the multi-systemic involvement of the virus, but its impact on disrupting the endocrine system--with more long-term effects--can also no longer be overlooked, particularly in the context of countries already struggling with prevalent endocrine diseases.^2^

In Pakistan, the prevalence of hyperthyroidism and subclinical hyperthyroidism is 5.1% and 5.8%, respectively, and hypothyroidism is at 4.1%. Even though hyperthyroid conditions are relatively rare in Pakistan.^4^ Our prevalence seems to be well above the prevalence of both disorders in Europe (0.7%) and the United States (0.5%), while in our closest neighbor, India, subclinical and overt hyperthyroidism were present in 1.6% and 1.3%, respectively and hypothyroidism is comparable at 3.4%.^9,11^

Unlike other countries where COVID-19 has decimated population bases, Pakistan seems to have emerged relatively unscathed from the pandemic.^12^ However, given the thyroid abnormalities which have emerged in our study sample, it would seem that the long-term effects of COVID-19, particularly with regards to follow-up of critically ill patients, may be overlooked especially for elderly patients, in which the symptoms of excessive TT3 in circulation—body aches, confusion, increased heart rate—may simply be dismissed as recovery from a viral infection. Concurrently, a thyroid storm with raised inflammatory markers also has overlapping signs and symptoms with the cytokine storm characteristic of COVID-19 worsening in critically ill patients.^8,13,14,15^ Indeed, precedent for the effect of a previous coronavirus affecting the thyroid was established by post-mortem studies of thyroid samples from patients infected with SARS during the 2002 outbreak.^16^ This immunogenic and hormonal overlap of a novel disease may further complicate the management and recovery of COVID-19, as corticosteroids used in the treatment of severe COVID-19 may inadvertently cause auto-immune damage to the thyroid gland.^3,16^

A study expressed a reduced TSH level in COVID-19 patients. It can be explained by two hypotheses.^17^ One is direct follicular damage by SARS-COV-2. This was contradicted by another study which proposed pituitary dysfunction instead of thyroid tissue destruction.^18^ However, there is a manifestation of thyroid dysfunction in critically ill patients, commonly called sick euthyroid. There are multiple mechanisms contributing to this condition, including alterations in TSH secretion, binding of thyroid hormone to transport proteins, and peripheral uptake of thyroid hormone. This condition is a pathologic response to acute illness and treatment with thyroid hormone imparts no benefit in sick euthyroid.^19^ At present, no study has investigated effect of COVID-19 on sick euthyroid. Our study cohort demonstrated significantly raised TSH and depressed TT3 levels in patients with COVID-19 pneumonia as compared to non-COVID-19 pneumonia with corresponding severity. Therefore, it is our understanding that COVID-19 affects thyroid functions and acute phase reactants irrespective of disease severity when compared with similar non-COVID-19 patients and sick euthyroidism is not present in this cohort.

This study had a number of limitations. One was a small cohort of consenting patients and less duration of follow-up. Second was that free T3, free T4, and other pituitary hormones were not assessed at admission and follow-up due to the retrospective nature of the study. Our institute being a tertiary care, it received a higher than moderately severe COVID-19 cases. Hence, patients with mild COVID-19 were not assessed for thyroid functions.

## Conclusion

This retrospective study provides solid evidence of the high risk of altered thyroid function after COVID-19 pneumonia. We believe our study represents an important direction of exploration of the true cost of the disease, where the overt crises of COVID-19 may mask an emerging crisis of long-term endocrine dysfunctions. Clinical vigilance must be practiced by physicians when monitoring for recovery from COVID-19 for signs and symptoms of thyroid hormone disruptions, particularly in patients of South Asian descent. Further prospective studies will clarify the clinical and prognostic relevance of abnormal behavior of the thyroid gland with COVID-19.

## Data Availability

Available upon request

## Acknowledgement

None

## Supporting document

Data in SPSS file available as supporting doc.

## References

1. Scappaticcio L, Pitoia F, Esposito K, Piccardo A, Trimboli P. Impact of COVID-19 on the thyroid gland: an update [published online ahead of print, 2020 Nov 25]. Rev Endocr Metab Disord. 2020;1–13. doi:10.1007/s11154-020-09615-z

2. Prompetchara E, Ketloy C, Palaga T. Immune responses in COVID-19 and potential vaccines: Lessons learned from SARS and MERS epidemic. Asian Pac J Allergy Immunol. 2020;38(1):1–9. doi:10.12932/AP-200220-0772

3. Wang W., Ye Y.X., Yao H. Evaluation and observation of serum thyroid hormone and parathyroid hormone in patients with severe acute respiratory syndrome. J Chin Antituberculous Assoc. 2003;25:232–234.

4. Khan A, Khan MMA, Akhtar S. Thyroid Disorders, Etiology and Prevalence. J of Medical Sciences. 2002; 2(2):89–94. doi: 10.3923/jms.2002.89.94

5. Chen M, Zhou W, Xu W. Thyroid Function Analysis in 50 Patients with COVID-19: A Retrospective Study. Thyroid. 2020 Jul 10. doi: 10.1089/thy.2020.0363. Epub ahead of print.

6. Lania A, Sandri MT, Cellini M, Mirani M, Lavezzi E, Mazziotti G. Thyrotoxicosis in patients with COVID-19: the THYRCOV study. Eur J Endocrinol. 2020 Oct;183(4):381–387. doi: 10.1530/EJE-20-0335. PMID: 32698147.

7. Clerkin KJ, Fried JA, Raikhelkar J, Sayer G, Griffin JM, Masoumi A, et al. COVID-19 and Cardiovascular Disease. Circulation. 2020 May 19;141(20):1648–1655. doi: 10.1161/CIRCULATIONAHA.120.046941.

8. Dworakowska D, Grossman AB. Thyroid disease in the time of COVID-19. Endocrine. 2020;68(3):471–474. doi:10.1007/s12020-020-02364-8

9. Taylor PN, Albrecht D, Scholz A, Gutierrez-Buey G, Lazarus JH, Dayan CM, Okosieme OE. Global epidemiology of hyperthyroidism and hypothyroidism. Nat Rev Endocrinol. 2018 May;14(5):301–316. doi: 10.1038/nrendo.2018.18.

10. Noor AU, Maqbool F, Bhatti ZA, Khan AU. Epidemiology of CoViD-19 Pandemic: Recovery and mortality ratio around the globe. Pak J Med Sci. 2020;36(COVID19-S4):S79–S84. doi:10.12669/pjms.36.COVID19-S4.2660

11. Unnikrishnan AG, Menon UV. Thyroid disorders in India: An epidemiological perspective. Indian J Endocrinol Metab. 2011;15(Suppl 2):S78–S81. doi:10.4103/2230-8210.83329

12. Abid K, Bari YA, Younas M, Tahir Javaid S, Imran A. Progress of COVID-19 Epidemic in Pakistan. Asia Pac J Public Health. 2020;32(4):154–156. doi:10.1177/1010539520927259

13. Dasgupta A, Kalhan A, Kalra S. Long term complications and rehabilitation of COVID-19 patients. J Pak Med Assoc. 2020;70(Suppl 3)(5):S131–S135. doi:10.5455/JPMA.32

14. Bellastella G, Maiorino MI, Esposito K. Endocrine complications of COVID-19: what happens to the thyroid and adrenal glands?. J Endocrinol Invest. 2020;43(8):1169–1170. doi:10.1007/s40618-020-01311-8

15. Brancatella A, Ricci D, Cappellani D, Viola N, Sgrò D, Santini F, Latrofa F. Is Subacute Thyroiditis an Underestimated Manifestation of SARS-CoV-2 Infection? Insights From a Case Series. J Clin Endocrinol Metab. 2020 Oct 1;105(10):dgaa537. doi: 10.1210/clinem/dgaa537.

16. Brancatella A, Ricci D, Viola N, Sgrò D, Santini F, Latrofa F. Subacute Thyroiditis After Sars-COV-2 Infection. J Clin Endocrinol Metab. 2020;105(7):dgaa276. doi:10.1210/clinem/dgaa276

17. Wang W, Ye YX, Yao H. Evaluation and observation of serum thyroid hormone and parathyroid hormone in patients with severe acute respiratory syndrome. J. Chin. Antituberculous Assoc. 2003; 25:232–234.

18. Leow MK, Kwek DS, Ng AW, Ong KC, Kaw GJ, Lee LS. Hypocortisolism in survivors of severe acute respiratory syndrome (SARS). Clin Endocrinol (Oxf). 2005; 63(2):197–202. doi: 10.1111/j.1365-2265.2005.02325.

19. Dworakowska D, Grossman AB. Thyroid disease in the time of COVID-19. Endocrine. 2020 Jun;68(3):471–474. doi: 10.1007/s12020-020-02364-8.

